# Investigation of Genetic Variants and Causal Biomarkers Associated with Brain Aging

**DOI:** 10.1101/2022.03.04.22271813

**Authors:** Jangho Kim, Junhyeong Lee, Seunggeun Lee

## Abstract

Delta age is a biomarker of brain aging that captures differences between the chronological age and the predicted biological brain age. Using multimodal data of brain MRI, genomics, and blood-based biomarkers and metabolomics in UK Biobank, this study investigates an explainable and causal basis of high delta age. A visual saliency map of brain regions showed that lower volumes in the fornix and the lower part of the thalamus are key predictors of high delta age. Genome-wide association analysis of the delta age using the SNP array data identified associated variants in gene regions such as KLF3-AS1 and STX1. Mendelian randomization (MR) for all metabolomic biomarkers and blood-related phenotypes showed that immune-related phenotypes have a causal impact on increasing delta age. Our analysis revealed regions in the brain that are susceptible to the aging process and provided evidence of the causal and genetic connections between immune responses and brain aging.

## Introduction

Aging is a primary risk factor for a myriad of health problems. Since aging proceeds at different rates for each individual, various methods to measure the biological age have been developed for a more accurate diagnosis of health status. Among the concerns related to aging, cerebral atrophy, which leads to cognitive decline, is a substantial risk to the individual well-being, constituting a major public health burden. Brain volume loss is also associated with neurodegenerative diseases such as Alzheimer’s disease and Parkinson’s disease [1, 2].

The aging process in different brain regions can be detected through structural and functional Magnetic Resonance Imaging (MRI). As large-scale datasets such as UK Biobank that contain neuroimaging data are becoming available, there have been efforts to accurately predict an individual’s chronological age with the neuroimaging datasets. Franke *et al*. [3] used principal component analysis and relevance vector machine to predict age. Studies since then primarily used neural network models for prediction and data-driven feature extraction [4-11]. The convolutional neural network (CNN) models have been used with a high level of accuracy. The mean absolute error of the prediction in most literature with CNN models is between 2.14 and 3.4 years.

The difference between the predicted age and the actual chronological age, called delta age, has been used as an aging biomarker [3, 4]. After estimating the delta age, phenome-wide and genome-wide association tests have been conducted to identify significantly associated genetic and clinical factors. Recent studies have shown that bone mineral density, blood pressure, and type 2 diabetes are associated with delta age [6, 7]. Genome-wide association analyses identified that KANSL1, MAPT-AS1, CRHR1, NSF in chromosome 17, KLF3 (chromosome 4), RUNX2 (chromosome 6), and NKX6-2 gene (chromosome 10) were significantly associated [5, 9, 10]. When combined with the cognitive test results, SNPs in MED8, COLEC10, and PLIN4 genes were also significantly associated [10].

To extend our understanding of the genetic and molecular basis of brain aging, we analyzed multimodal UK Biobank data. Compared to the previous studies, our analysis includes whole-exome sequencing (WES) and metabolomics data, which enabled us to identify novel genetic and biomarker associations. In addition, we carried out large-scale Mendelian randomization studies of 310 blood and metabolomic phenotypes to identify causal biomarkers and used an explainable AI method for medical images to identify brain regions that drive high delta age.

## Results

### Overview of the Analysis

Figure 1 provides an overview of our analysis. First, a 3D CNN model was trained for age prediction with the T1-weighted structural brain MRI of healthy white British samples in the UK Biobank, excluding individuals with diseases related to cancer, diabetes, dementia, and mental disorders. The training was conducted via cross-validation to use all available samples in the downstream analysis (see Methods). The Integrated Gradients (IG) method was then used to identify an accurate attribution of each voxel (volume + pixel) to the prediction [12]. Second, genome-wide association tests were conducted on different test levels to uncover novel loci associated with brain aging. We applied a single-variant test, SAIGE (Scalable and Accurate Implementation of GEneralized mixed model), to the array-genotyped and imputed markers, and a gene-based test, SAIGE-GENE+, to the WES datasets [13, 14]. Lastly, we examined linear and nonlinear causal relationships between the delta age and the phenotypes (metabolomics and blood) with Mendelian randomization methods. The number of samples used in each step is in Table 1.

**Table 1.** The number of samples in each stage of the analysis

| 3D CNN<br>(Training) | 3D CNN<br>(Test) | IG | GWAS<br>(SNPs) | GWAS<br>(WES) | MR and<br>Association<br>(Metabolo<br>mics) | MR and<br>Associatio<br>n (Blood<br>Chemistry) |
| --- | --- | --- | --- | --- | --- | --- |
| 25,656 | 34,129 | 34,129 | 34,129 | 30,812 | 8,464 | 34,129 |

**Figure 1.**
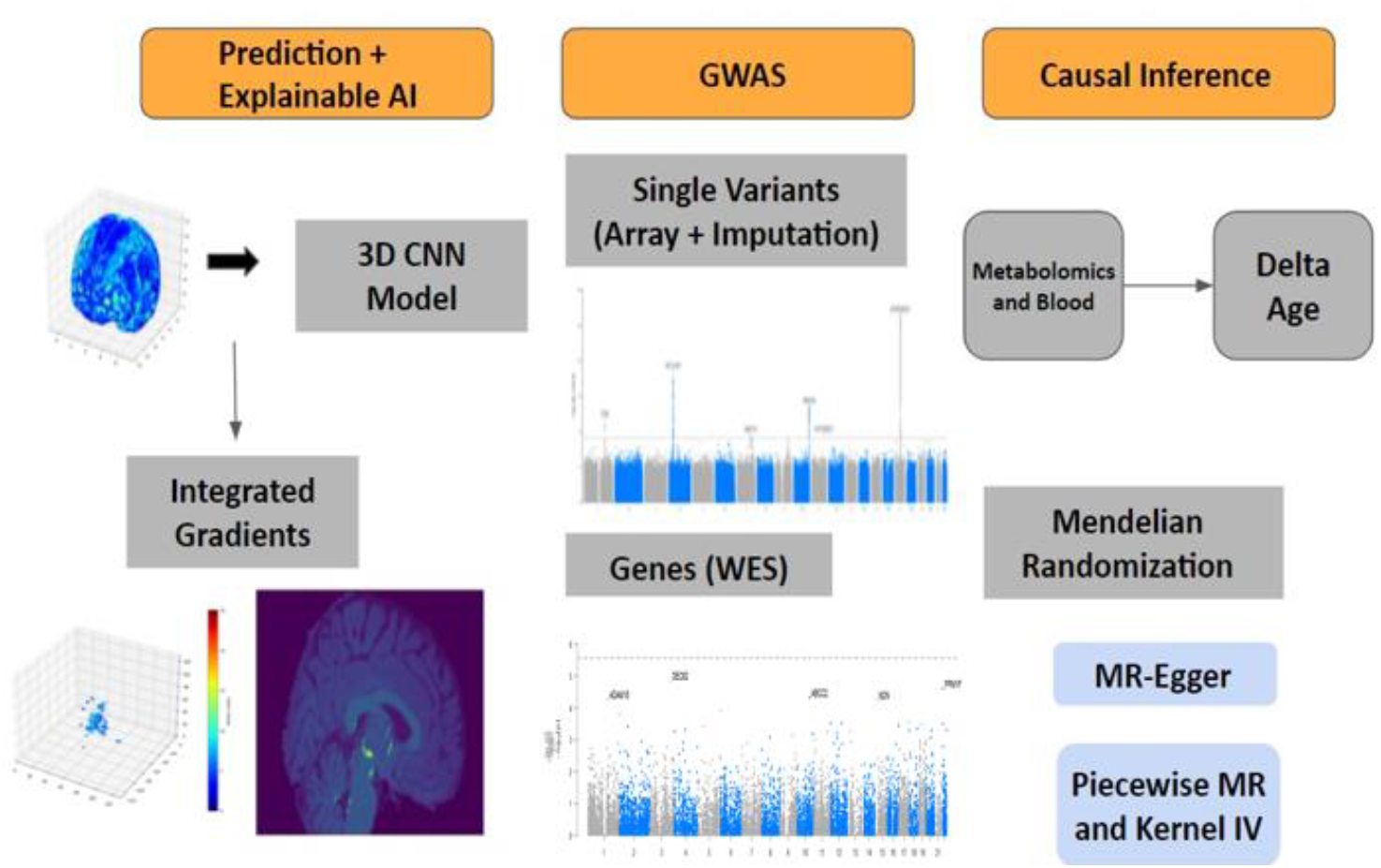
Overview of the analysis

### Age Prediction Accuracy and Saliency Map

Table 2 and Figure 2 show the prediction results of the 3D CNN model. We used the cross-validation scheme to use all available samples [9]. Table 2 shows the mean absolute error of the samples with diseases and the samples without diseases and four-fold cross-validation groups (see Methods). The mean absolute error in the healthy individuals was about 2.6406 years in the test set and 0.8989 years in the training set. The existing studies on brain age estimation had similar accuracy to this result [6-9]. The mean absolute error in the samples with diseases was 2.651 years. Figure 2 (a) shows the strong positive correlation between the chronological age (x-axis) and the predicted age (y-axis). The age-related bias was corrected by linear regression on the chronological age (Figure 2 (b)) [15]. Figure 2 (c) is a scatter plot between the chronological age and the adjusted delta age.

**Table 2.**
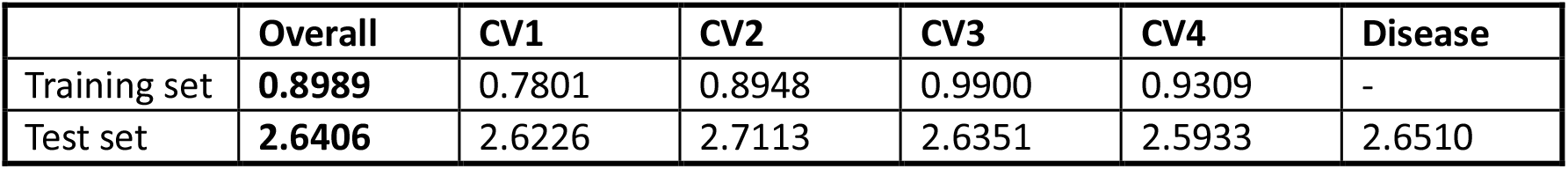
The mean absolute error (MAE) of individuals without diseases, each cross-validation batch, and individuals with diseases

|  | Overall | CV1 | CV2 | CV3 | CV4 | Disease |
| --- | --- | --- | --- | --- | --- | --- |
| Training set | **0.8989** | 0.7801 | 0.8948 | 0.9900 | 0.9309 | - |
| Test set | **2.6406** | 2.6226 | 2.7113 | 2.6351 | 2.5933 | 2.6510 |

**Figure 2.**
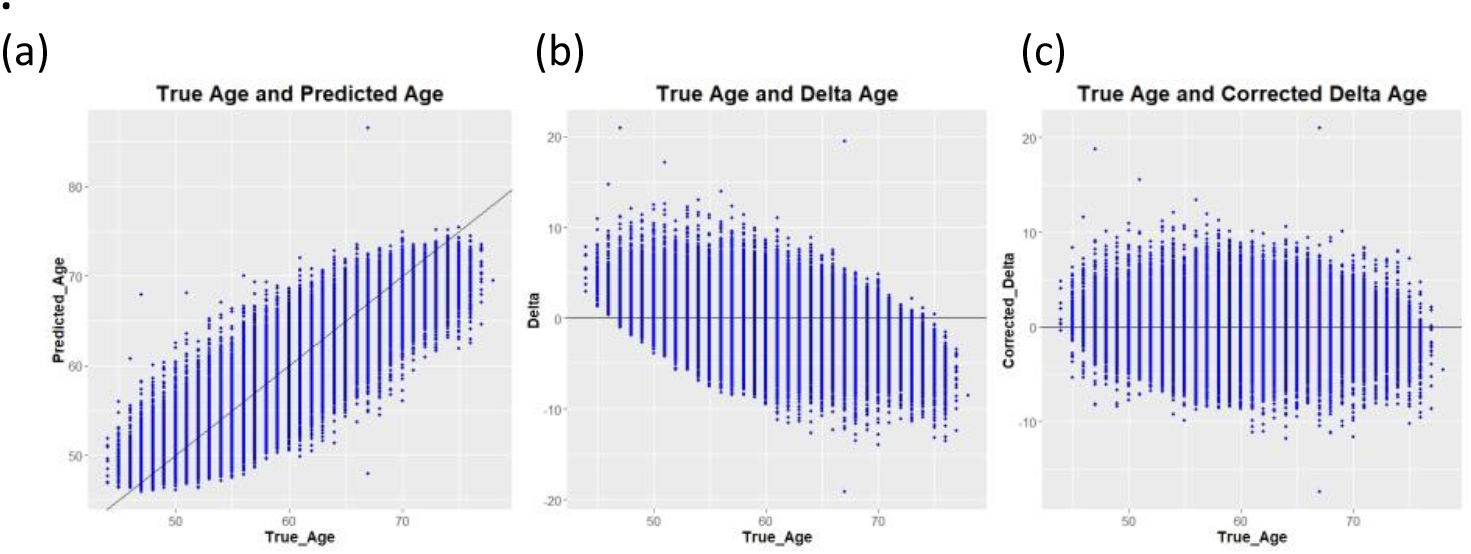
Scatterplots of chronological age (x-axis) vs. (a) predicted age, (b) delta age, and (c) bias-adjusted delta age (y-axis)

Figure 3 shows the saliency maps of the age prediction model with integrated gradients. Absolute values of the integrated gradients were averaged for 100 samples with the youngest predicted age. Figure 3 (a) shows the voxels with averaged integrated gradients greater than five. According to the Automated Anatomical Labeling atlas (AAL) and the Natbrainlab atlas, when highlighting the regions with integrated gradients greater than ten, the regions were the fornix and the lower part of the thalamus (Figure 3 (b)) [16, 17]. Similar results were shown when 100 samples were chosen randomly or by descending order of the delta age values (data not shown).

**Figure 3.**
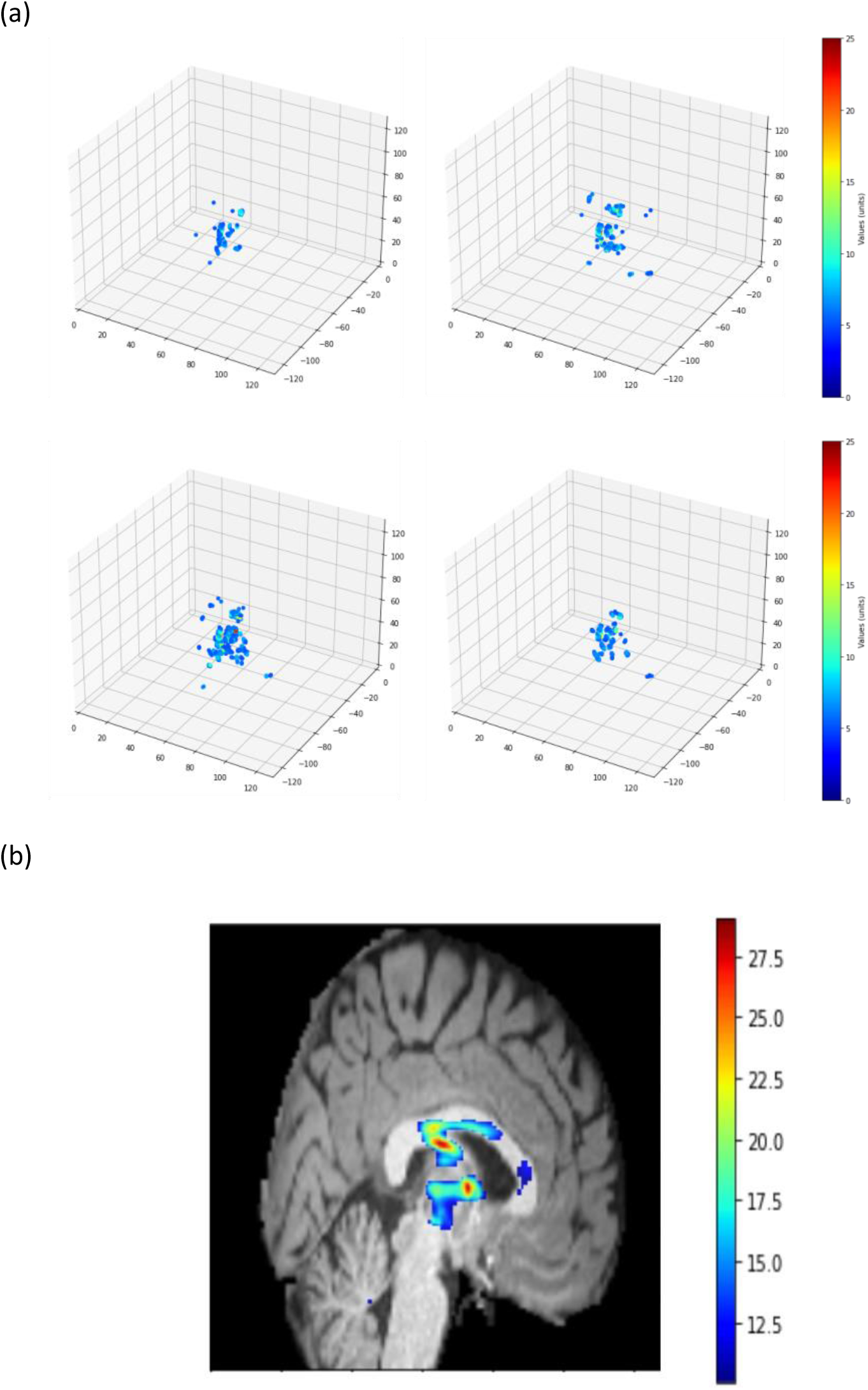
Integrated gradient (IG) analysis results. (a) Points with absolute IG > 5 in the four CV model sets from 100 individuals with the youngest predicted age. (b) A brain image at the x-coordinate of 62. Voxels with absolute IG > 10 are highlighted with colors.

### Genetic Variants Associated with Delta Age

We performed genetic association analyses for the predicted delta age. We carried out single-variant tests for 38 million array-genotyped and imputed genetic variants using SAIGE. Since the single-variant test for rare variants in WES has low power, we used a gene-based test method for the WES dataset [18]. We used SAIGE-GENE+ to test for rare variant associations (18,308 genes).

Figure 4 is the Manhattan plot of single-variant analysis results from array-genotyped and imputed data. Table 3 lists significant variants. The single-variant test results showed that five loci in chromosomes 1, 4, 6, 10, 11, and 17 were significantly associated with the delta age. The nearest genes were STX6, MR1, KLF3-AS1, WNT16, INPP5A, NKX6-2, and several genes in chromosome 17, including KANSL1, MAPT-AS1, and NSF. Genetic heritability calculated by the variance components in the SAIGE model was 21.5 percent.

**Table 3.** Significant loci associated with delta age. Single-variant test, SAIGE, was applied to array-genotyped and imputed genetics data.

| Chromo<br>some | Location<br>(GRCh37) | rsID | Nearest<br>gene | Effect<br>size | SE | P-value | MAF |
| --- | --- | --- | --- | --- | --- | --- | --- |
| 17 | 44,335,635 | rs147431626 | AC0058<br>29.2 | 0.0875 | 0.0091 | 5.36E-22 | 0.2272 |
| 4 | 38,638,966 | rs201791735 | KLF3-AS1 | 0.0661 | 0.0082 | 1.14E-15 | 0.3417 |
| 10 | 134,559,486 | -(deletion) | INPP5A | 0.0607 | 0.0089 | 1.10E-11 | 0.2401 |
| 1 | 180,963,000 | rs534102361 | STX6 | -0.0480 | 0.0076 | 3.46E-10 | 0.5714 |
| 11 | 112,766,643 | rs536165403 | AC0169<br>02.1 | 1.7857 | 0.3163 | 1.64E-08 | 0.0002 |
| 7 | 120,981,641 | -(deletion) | WNT16 | -0.0438 | 0.0078 | 1.90E-08 | 0.4698 |

**Figure 4.**
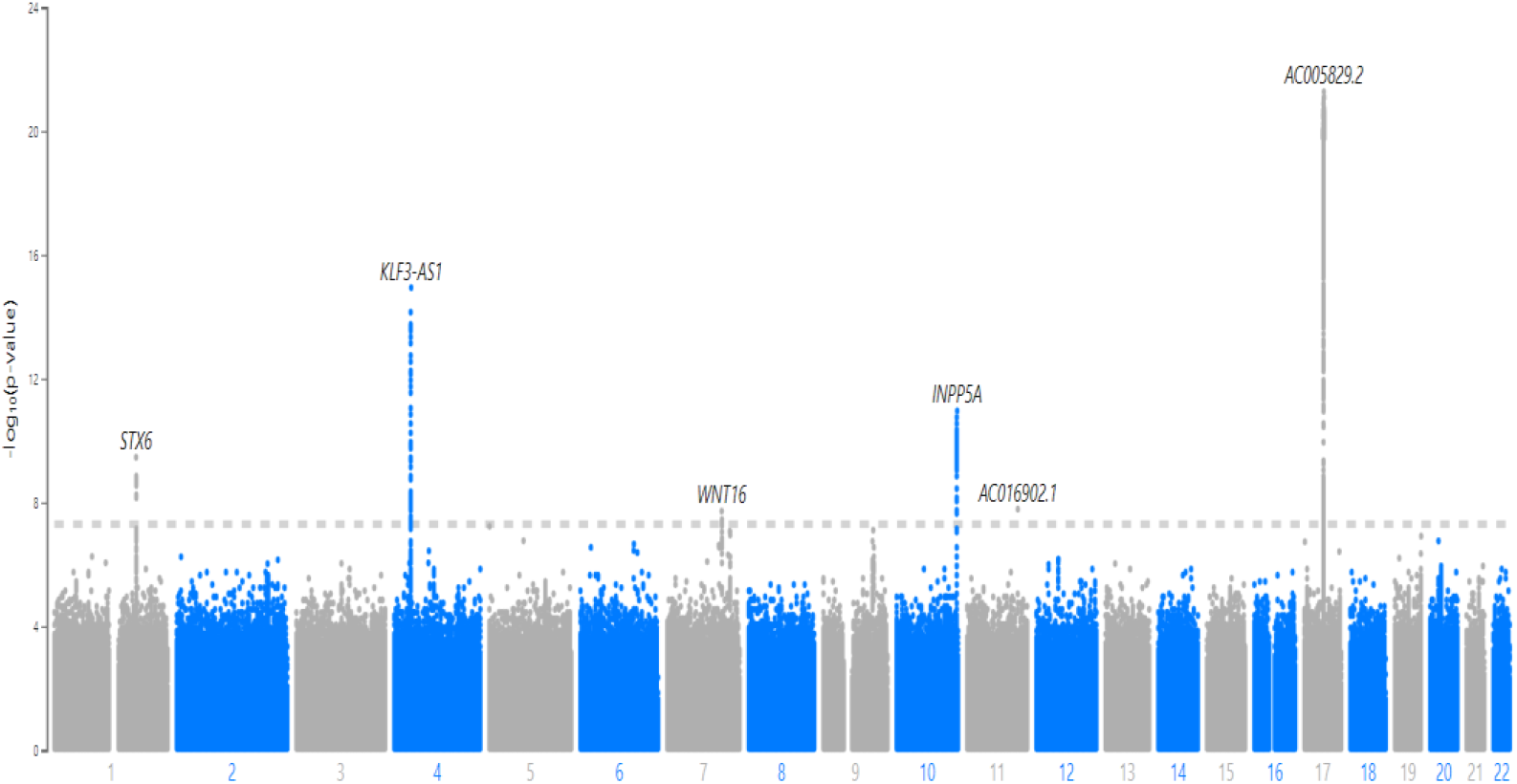
Manhattan plot of Single-variant tests with the array-genotyped and imputed genetics data on the delta age.

The gene-based rare variant test on the WES dataset identified no significant genes. The p-value threshold was the Bonferroni corrected level of 0.05 (0.05/18,308). SEC62, PPM1F, ABCC2, ADMA15, and NDN were the top five genes with the smallest p-value. (Table 4)

**Table 4.** Top 5 genes with the smallest p-value. Gene-based test, SAIGE-GENE+, was applied to the WES data. P-values were calculated using Cauchy-combination in SAIGE-GENE+.

| Chromosome | Starting location (GRCh38) | Gene | P-value |
| --- | --- | --- | --- |
| 3 | 169966635 | SEC62 | 1.38E-05 |
| 22 | 21919425 | PPM1F | 2.50E-05 |
| 10 | 99782640 | ABCC2 | 4.82E-05 |
| 1 | 155050566 | ADAM15 | 5.48E-05 |
| 15 | 23685400 | NDN | 5.64E-05 |

To check whether the delta age prediction was truly driven by voxel values in the fornix and the lower part of the thalamus, we carried out the same GWAS procedure with the average voxel value of the two regions. The SAIGE results showed that the significant loci associated with the two regions were also concentrated on chromosome 17 (Supplementary Table 1 and Supplementary Figure 2 (a), (b)). SLC39A8 and C16orf95 genes were commonly shown to be associated with the two regions. We also calculated the genetic correlation among delta age and average voxel values of the two regions and observed high genetic correlation values (Supplementary Table 2). Our analysis results clearly demonstrated the shared genetic basis of delta age and the two regions.

Additional validation on the delta age of 1,610 healthy non-British white samples was conducted. In single-SNP GWAS, two SNPs in chromosome 4 with no specific gene region and two SNPs in chromosome 17 (rs375822897 in PLEKHM1 and rs56303031 in LINC02210-CRHR1) had p-values less than 0.05. Due to the small sample size (905 samples), none of the genes had p-values < 0.05 in non-British white samples.

### Causal Biomarkers of Delta Age

Among the 310 phenotypes (249 metabolomic phenotypes and 61 phenotypes related to blood), 59 had p-values less than 0.05 in causal estimates from at least one of the three linear MR methods (MR-Egger regression, inverse variance weighting, and weighted median). Table 5 lists the top five phenotypes by MR-Egger regression (Eosinophil count, Eosinophil percentage, Neutrophil count, Total protein, and White blood cell count), and Supplementary Table 3 shows the other 54 phenotypes. The Top five phenotypes were immune-related biomarkers and had positive relationship with the delta age. Among them Eosinophil count (P-value=5.16E-06) was statistically significant after the Benjamini-Hochberg procedure (FDR=0.05).

**Table 5.** Top 5 blood biomarkers with the smallest p-value. Causal estimates, standard error, and p-values of blood biomarkers in the MR-Egger regression.

| Biomarker | Estimates | SE | P-value |
| --- | --- | --- | --- |
| Eosinophil count | 0.151 | 0.033 | 5.16E-06 |
| Eosinophil percentage | 0.156 | 0.045 | 5.04E-04 |
| Neutrophil count | 0.153 | 0.048 | 1.28E-03 |
| Total protein | 0.181 | 0.057 | 1.42E-03 |
| White blood cell (leukocyte) count | 0.100 | 0.038 | 8.01E-03 |

Figure 5 is a PheWAS plot of the causal estimates from the MR-Egger regression. Overall, phenotypes related to white blood cells showed more significant causal relationships with delta age than other phenotypes. Similar results were replicated by the weighted median method (Supplementary Figure 3 and Supplementary Table 4).

**Figure 5.**
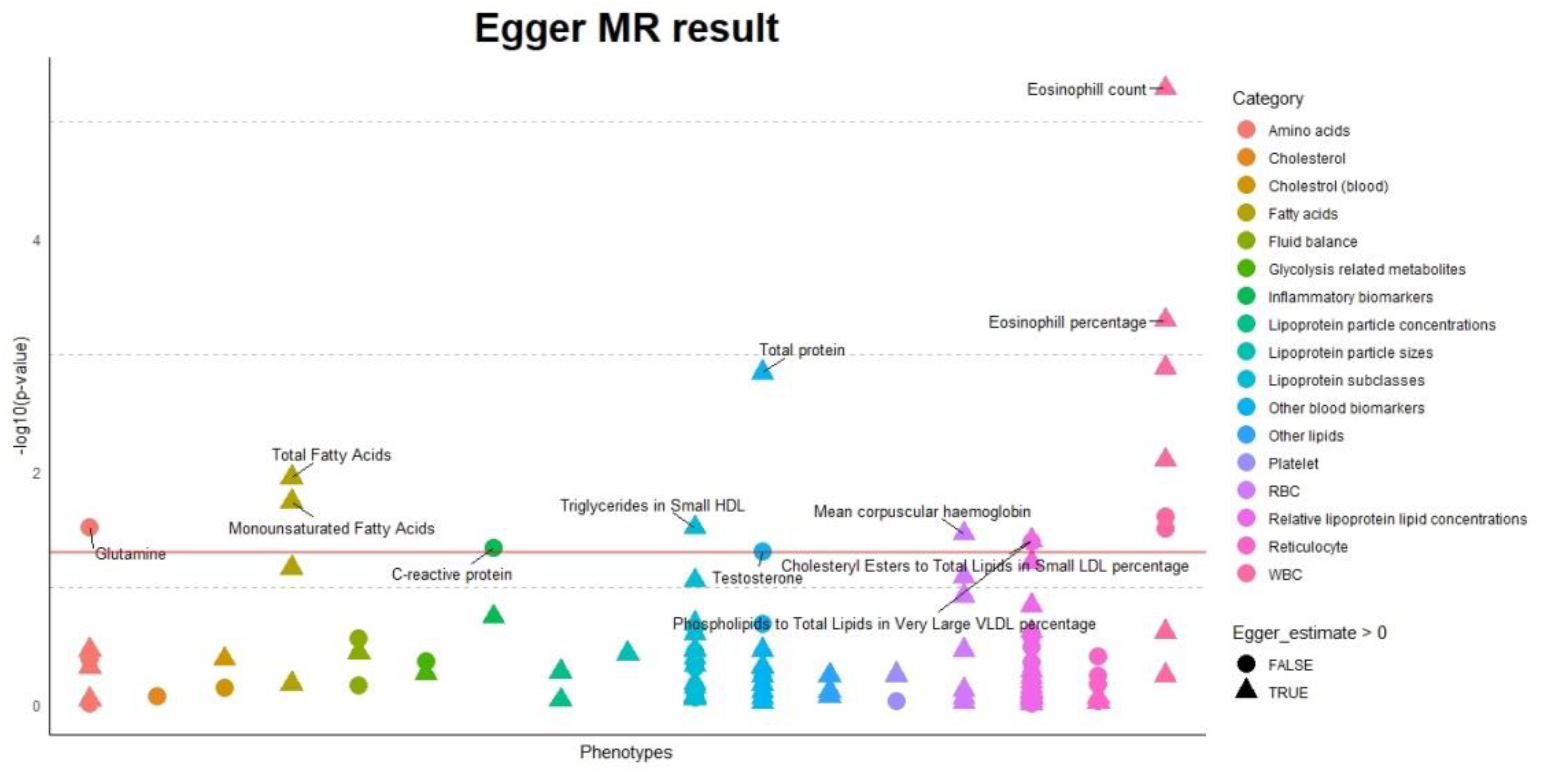
A PheWAS plot of Linear MR causal estimates from MR-Egger regression. The horizontal red line is the p-value threshold of 0.05. Each color indicates a different group of biomarkers. The two most significant in each group with p-values < 0.05 were labeled. The shape of the points indicates the direction of the causal effect. Triangles are the biomarkers that contribute to higher delta age, and circles are the biomarkers that contribute to lower delta age.

We also carried out association analysis with a linear regression model. HbA1C (P-value=7.31E-28) and Glucose (P-value=1.11E-26) were most significantly associated with the delta age (Supplementary Figure 4 and Supplementary Table 5) with a positive association direction. This may indicate the association between diabetes and brain aging.

Nonlinear MR analysis using piecewise MR and kernel IV showed similar results. Total cholines (P-value=0.03413), total lipids in small LDL (P-value=0.03599), and cholesteryl esters to total lipids in very large HDL percentage (P-value=0.01002) passed the test of the assumptions for instrument variable regression and returned p-value less than 0.05 in the trend test, indicating that there was nonlinearity in the causal relationship between the biomarkers and delta age (the values in the parenthesis indicate the p-values). Total cholines showed a causal relationship with the threshold. It increases delta age when it is above 2.5 mmol/l. The other two biomarkers showed an inverted U-shaped relationship (Supplementary Figure 5). However, when applying multiple testing corrections, none of the three biomarkers were significant.

## Discussion

In this paper, we have analyzed the risk factors of brain aging with multimodal data. The CNN model that predicts age from brain MRI had high accuracy with a mean absolute error of 2.64 years. Visual information of the regional importance in the brain was extracted from the neural network model. Genetic variants and biomarkers that have significant links to brain aging were identified using GWAS methods and Mendelian randomization.

We used integrated gradients to make accurate saliency maps by incorporating information from a wider range of pixel values not present in the original images. This information revealed important brain regions missed by other mapping methods. The saliency map in the previous studies highlighted the brain regions such as the hippocampus, brainstem, and amygdala [6, 7, 19]. When highlighting the important points with higher integrated gradients in our study, they were centered on the fornix and the lower part of the thalamus. This indicates that the aging process affects the brain mainly through the atrophy in the inner area connected to memory and learning ability [20-23]. In addition, genetic variants associated with volumes of these regions and delta age were highly similar, supporting the result.

Investigating gene-level rare variant associations in the WES data, we identified no significant genes associated with the delta age. However, the NDN gene, one of the top five genes with the smallest p-value, is known to play an important role in neural differentiation and survival of postmitotic neurons [24, 25]. In addition, this gene is associated with the PWS (Prader-Willi syndrome), which is known to have a significantly higher delta age in the PWS [26].

In the single-variant test of array-genotyped and imputed variants, we replicated strong association signals in chromosome 17. The significant variants in other chromosomes were in STX6, MR1, KLF3-AS1, WNT16, INPP5A, and NKX6-2. STX6 and KLF3-AS1 relate to carcinogenesis [27, 28]. MR1 and WNT16 participate in immune response via antigen presentation to T cells and lymphocyte proliferation [29, 30]. Mutations in INPP5A and NKX6-2 were shown to cause neurologic problems [31, 32].

Our investigation of the causal effects of 310 blood and metabolomic biomarkers on delta age showed the potential causal roles of immune responses to delta age. Especially, biomarkers related to white blood cells had a significant causal effect. This claim is supported by existing literature [33, 34].

This study, however, is subject to several limitations. First, the analysis was done in the European ancestry group only. The aging process and genetic variants associated with aging can differ according to ancestry. Second, replication of the results in an independent dataset was not conducted due to a lack of datasets with genetics data, extensive biomarkers, and brain MRI. Future research should focus on addressing the limits and making results more generalizable.

In conclusion, our multimodal data analysis shows many aspects of brain aging, including brain regions most affected by the aging, associate genes, and causal biomarkers. As more biobanks with multimodal data are collected, more diverse aspects of brain aging can be revealed. Biobanks of other ancestry groups can identify novel biomarkers associated with brain aging not identified in this study. In addition, potential mediating factors between immune responses and brain aging can be revealed. This would allow a deeper understanding of brain aging mechanisms that develop proper prevention and treatment.

## Methods

### Data Preprocessing

T1-weighted structural MRI images of 34,129 white British samples in the UK Biobank were used for the analysis to minimize the effect of ancestry (average age of 60.964) [35]. All of the images downloaded from the UK Biobank had been normalized into MNI152 space (Montreal Neurosciences Institute) to render the comparison of each voxel possible [36]. The samples were selected if they had proper images and if they had no relation to any other individuals in the dataset. Each image was resized from 182×216×182 to 128×128×128 to reduce the computation cost. First, a part of z-axis voxels (from 26 to 153 out of 182 points) was selected to include various brain regions in the prediction task. The voxels in the upper outermost surface of the brain were excluded since they were considered negligible in the prediction and redundant due to the inclusion of other parts of the cerebral cortex. Second, each 2-dimensional 182×216 image (x, y-axis) in z-axis points was resized to a 128×128 image with the nearest neighbors scaling algorithm. The preprocessing of nifti format MRI images was performed with oro.nifti and OpenImageR packages in R [37, 38].

For the genetics data, we downloaded bgen files of 93 million array-genotyped and imputed variants dataset. And we used plink files of 26 million whole-exome sequencing (WES) variants dataset from the UK Biobank. The 450K WES data were used in our analysis, and the analysis was performed on the DNA nexus.

The 249 metabolomic phenotypes and 61 biomarkers from blood assays and blood count were used in the Mendelian randomization (MR) analysis. The metabolomic phenotypes and biomarkers were collected separately from MRI imaging, from 2006 to 2010; this would enable the investigation of the biomarkers’ longitudinal and cumulative effect on the brain. The missing values in the selected biomarkers were imputed with multiple imputation by chained equations (MICE) to fit the missing values to the overall multivariate distribution [39]. After filling the missing values, 8,464 individuals with the brain MRI had corresponding values of metabolomic phenotypes, and all 34,129 individuals had values of blood-related phenotypes. The 310 selected biomarkers were divided into 17 groups, including amino acids, cholesterol, and glycolysis-related metabolites.

### 3D CNN Prediction Model and Integrated Gradients

3D CNN model was used to predict the age of the individuals. Supplementary Figure 6 is the overall network structure of the prediction model. The neural network model takes the resized images (128×128×128) as input and has less than three million parameters to train. The spatial dropout layers were added in the first two blocks to prevent the model from overfitting. The kernel size is 3×3×3 in convolution layers and 2×2×2 in pooling layers. The model conducts max-pooling until the size of an image in each feature becomes 2×2×2. The number of features increases as the input image size reduces. Adam optimizer with learning rate 0.001 and He uniform initializer were used since the activation function is the rectified linear unit (ReLU).

Healthy white British individuals (25,656) were selected to train the prediction model. Individuals with diseases (all types of cancers, diabetes, neoplasm, dementia, and mental disorders) were excluded from the training process. The dataset with healthy individuals was divided into four sets (CV1, CV2, CV3, and CV4) for four-fold cross-validation so that every sample is included in the test set at least once and has a predicted age value. When CV1 is the test dataset, the other three sets become the training dataset. For each training set, three separate models (the same structure in Supplementary Figure 6 with different initial weights and dropouts) were trained for more robust prediction. They constitute a single model set. After training the models, the test images were given to the models as input. The average of the predictions from the three models becomes the final predicted age of the test images, hence a total of 12 models to train (three models for each of the four cross-validation batches). Prediction of the age of individuals with diseases was made with the average of the predicted age from the four trained model sets. The delta age value of each sample was calculated by subtracting the individual’s chronological age from the predicted age. The age-related bias in the delta age value was adjusted through linear regression on the chronological age. The adjusted delta age values were used in the later association tests.

To identify which regions in the brain contributes significantly to age prediction, the Integrated Gradients (IG) method was used. The IG method is an explainable AI method for neural networks that uses multiple images between blank and original images [12]. In this study, the number of images generated for each sample was 101 in reference to the recommended step size in the original paper.

### Genome-wide Association Test with Single Variants and Gene Regions

We used SAIGE for array-genotyped and imputed variants and SAIGE-GENE+ for the WES variants. Variants with minor allele frequency less than 0.0001 were excluded in the SAIGE analysis. SAIGE uses a mixed effect model to account for the relatedness among the individuals. SAIGE-GENE+ is a gene-based rare variant association test and it performs BURDEN, SKAT, and SKAT-O tests [14]. Since the number of tests decreases in the gene-based test, multiple testing correction is less stringent.

34,129 (=*N*) samples were used in SAIGE analysis. The delta age values of the individuals were inverse-normal transformed. Covariates were sex, age, ten principal component scores, and four dummy variables which indicate different cross-validation test sets plus samples with diseases. *N* × *N* genomic relation matrix (GRM) was calculated 784,256 markers in called autosomal genotypes. Leave-one-chromosome-out (LOCO) option was applied when estimating the GRM.

18,308 regions were identified with annotation on the WES genotype by the ANNOVAR software [40]. The size of the samples *n* in the gene-based test was 30,812 (white British individuals with proper MRI images included in the UK Biobank 450k whole-exome sequencing data). The SAIGE-GENE+ analysis was done with 3 MAF cutoffs (MAF=0.01, 0.001, 0.0001) and 3 functional annotation groups (LOF, LOF+Missense, LOF+Missense+Synonymous). In addition to the same covariates in the SAIGE test, the batch indicator variable was included to adjust for possible batch effect in the WES dataset.

Genetic correlation among delta age and the average volume of two brain regions (the fornix and the lower part of the thalamus) was derived using LD score regression with western European LD scores [41].

### Linear and Nonlinear Mendelian Randomization

We used variations of Mendelian randomization methods to identify the causal effect of biomarkers (exposure) on delta age (outcome). The overall workflow of the Mendelian randomization in this study is in Supplementary Figure 7. Instrument genetic markers for each of the 310 biomarkers were selected as follows. First, we chose variants significantly associated with the exposure (p-value under 5e-8) among the called autosomal genetic markers. The markers with minor allele frequencies less than 0.01 were pruned because the estimation of the effect size from rare variants is unstable. The GWAS summary statistics for the metabolomics measured by the Nightingale Health are from open datasets in MRC Integrative Epidemiology Unit at the University of Bristol (IEU) [42]. The GWAS results of blood phenotypes are from Pan-UK Biobank GWAS summary statistics by the Broad Institute (available at https://pan.ukbb.broadinstitute.org). We used effect size, standard error, and p-value from the samples with European ancestry. Second, the linkage disequilibrium (LD) pruning process was conducted with PLINK software with a window size of 50 base pairs to ensure that the selected instruments were independent of each other [44]. Pairs of variants with a correlation coefficient larger than 0.01 were LD pruned. Lastly, since the markers should not directly affect the outcome, variants with p-value with regard to the outcome less than 0.05 divided by the number of markers left were excluded. The effect size and standard error of the remaining markers were used in the MR analysis.

### Linear and Nonlinear Exposure-Outcome Relationship

We first calculated the causal impacts of the biomarkers (exposure) on delta age (outcome) with three linear Mendelian randomization methods: MR-Egger regression, weighted median method (WM), and inverse variance weighting method (IVW) [45-46]. The MendelianRandomization package in R was used to carry out the MR-Egger regression [47]. The estimates and p-values from the default version of MR-Egger regression were selected. The same was done for the estimates of the WM and IVW method. The estimates in all methods were assumed to follow the normal distribution. The Benjamini-Hochberg procedure was applied to control the false discovery rates at 0.05 [48].

We also carried out an association analysis between delta-age and blood-chemistry and metabolomics biomarkers. The following linear regression model of a single biomarker was used.

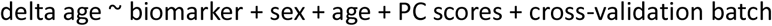

Nonlinear MR was conducted with the nlmr package in R [49]. The key rationale for using the nonlinear Mendelian randomization method is to find a nonlinear pattern of causal estimates from different ranges of exposure, as the impact of exposure on delta age can vary according to the ranges.

The assumptions for instrument variable regression were first thoroughly tested. Among the instruments selected for each biomarker from the linear MR procedure, the ones with positive effect sizes were collected. We constructed each sample’s single allele score *G* with those markers. Each marker has genotype 0, 1, or 2 for each sample. When there are *n* samples and *m* genetic markers, the allele score of sample *i* is in (1).

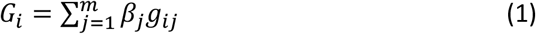

Here *β*_*j*_ is the effect size of the *j*th marker and *g*_*ij*_ is the genotype of sample *i* in the *j*th marker. The allele scores and the exposure values were tested for a significant positive relationship (p-value of the Pearson’s correlation coefficient lower than 0.05 using the *cor.test* function in R). Exclusion restriction was assumed to be testable with conditional independence tests (*Y* and *G* conditionally independent given *X*) because the directions of effect between *G, X*, and *Y* were fixed (*G* → *X* → *Y*). The allele score shows a genetically determined level of the biomarker, and the delta age was calculated from images taken after the level of the biomarker had been measured. In order to test for nonlinear conditional independence, Randomized Conditional Independence Test (RCIT) was used [50]. We repeated the RCIT three times. The assumptions were considered to have been met if none of the three tests had a p-value less than 0.05 with the null hypothesis of the conditional independence between *Y* and *G* given *X*. Only 12 out of 310 variables were found to satisfy all the assumptions for instrument variable regression.

Then, we conducted the piecewise MR analysis for the 12 variables that met the assumptions [49]. The samples were divided into ten groups according to deciles by the IV-free exposure. Assumptions of the IV-exposure relationship in the piecewise MR are the homogeneity and the linearity across all samples. These assumptions were tested by the heterogeneity test using Q statistics. If the null hypothesis of homogeneity in the estimates between the groups was not rejected in the heterogeneity test, the trend test was conducted on the biomarker. The trend test evaluated whether the local average causal effect in each group is explained by the average value of the exposure in the corresponding group. Kernel IV regression with radial basis function kernel was performed with the 12 passed phenotypes to check if the results were replicated. Due to the heavy computation cost, we sampled 2,000 individuals for the Kernel IV regression. The test values were 1,000 numbers with an equal distance between the minimum and the maximum of the exposure values.

## Data Availability

All data produced in the present study are available upon reasonable request to the authors. UK Biobank data were accessed under the accession number UKB: 45227. The summary statistics used in the Mendelian randomization analysis are available for public download at the IEU OpenGWAS Project (https://gwas.mrcieu.ac.uk) and the Pan-UK Biobank (https://pan.ukbb.broadinstitute.org).

https://pan.ukbb.broadinstitute.or

## Code Availability

The code used in the analyses is available at our Github page.

https://github.com/Flumenlucidum/Brain-Aging.

We used publicly available software for the analyses.

MICE: https://github.com/amices/mice

TensorFlow: https://www.tensorflow.org

SAIGE: https://github.com/weizhouUMICH/SAIGE

SAIGE-GENE+: https://saigegit.github.io//SAIGE-doc/

SKAT: https://github.com/leelabsg/SKAT

LDSC: https://github.com/bulik/ldsc

PLINK2: https://www.cog-genomics.org/plink/2.0

MendelianRandomization: https://github.com/cran/MendelianRandomization

nlmr: https://github.com/jrs95/nlmr

## Acknowledgements

This research was supported by Big Brain Project through the National Research Foundation of Korea (NRF) funded by the Ministry of Science and ICT (No. 2021M3E5D2A0102249311), and the Brain Pool Plus (BP+, Brain Pool+) Program through the National Research Foundation of Korea (NRF) funded by the Ministry of Science and ICT (2020H1D3A2A03100666). UK Biobank data were accessed under the accession number UKB: 45227.

## Author Contributions

J.K. and S.L. designed the experiments. J.K., J.L., and S.L. constructed and developed the age prediction model. J.K. and S.L. wrote the manuscript. All authors reviewed and approved the final version of the manuscript.

## Notes

### Competing Interest Statement

The authors have declared no competing interest.

### Clinical Protocols

https://github.com/Flumenlucidum/Brain-Aging

### Author Declarations

Ethics committee/IRB of Seoul National University gave ethical approval for this work.

### Summary of Updates

Association analysis plot was renewed.

