## Supplementary Figures for "Investigation of Genetic Variants and Causal Biomarkers Associated with Brain Aging"

### Supplementary Materials

(a)

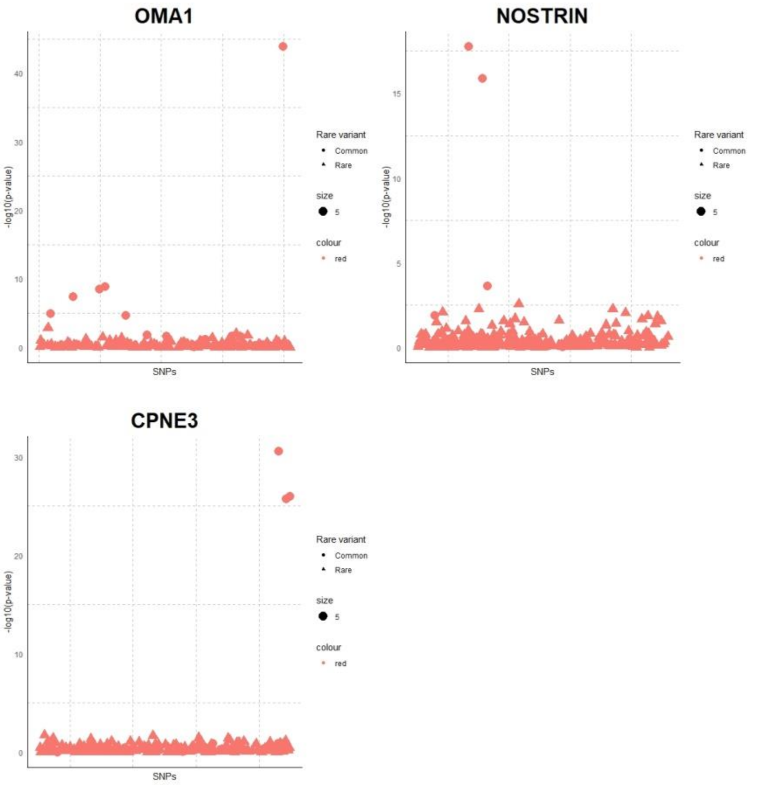


(b)

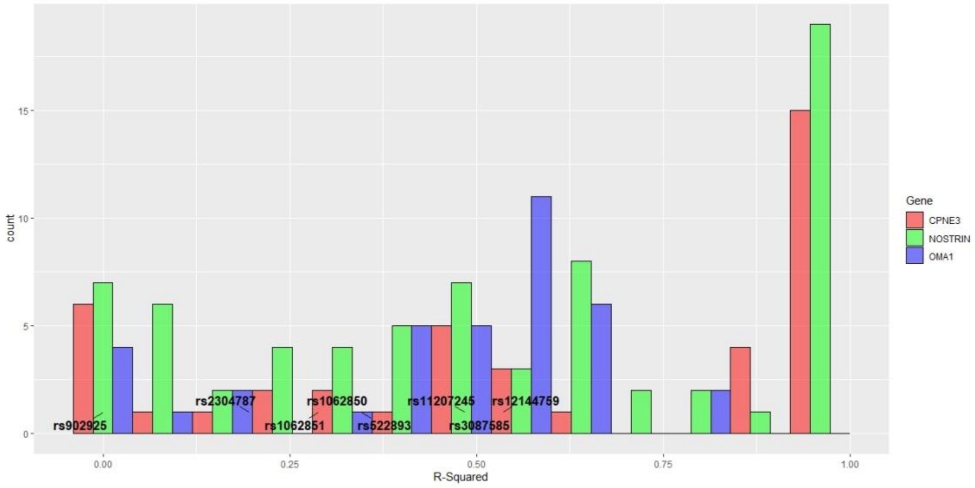


**Supplementary Figure 1** (a) SNP-level significance of OMA1, NOSTRIN, and CPNE3 genes) (b) Distribution of R-squared (square of correlation) between Array+Imputation SNPs and WES. Leading SNPs in the gene-based test are labeled with rsIDs. They have low R-squared values.

(a) SAIGE result on the fornix volume (single-variant test with array-genotyped and imputed data)

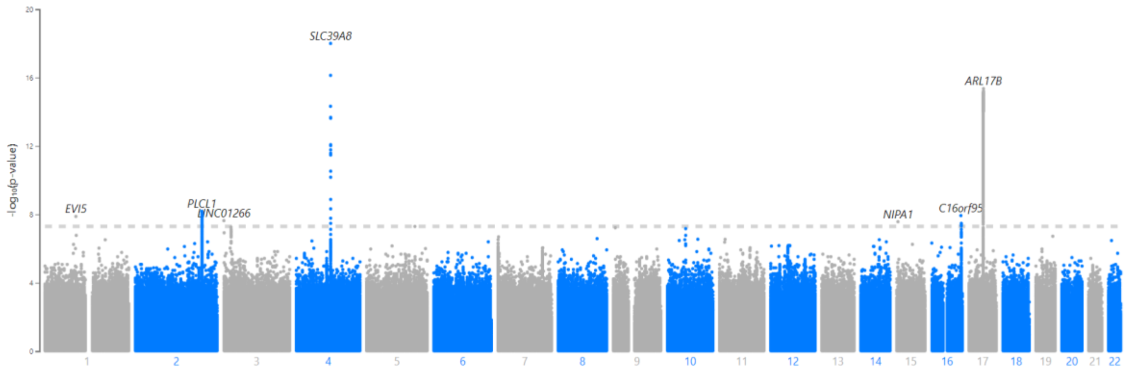


(b) SAIGE result on volume in the lower part of the thalamus (single-variant test with array-genotyped and imputed data)

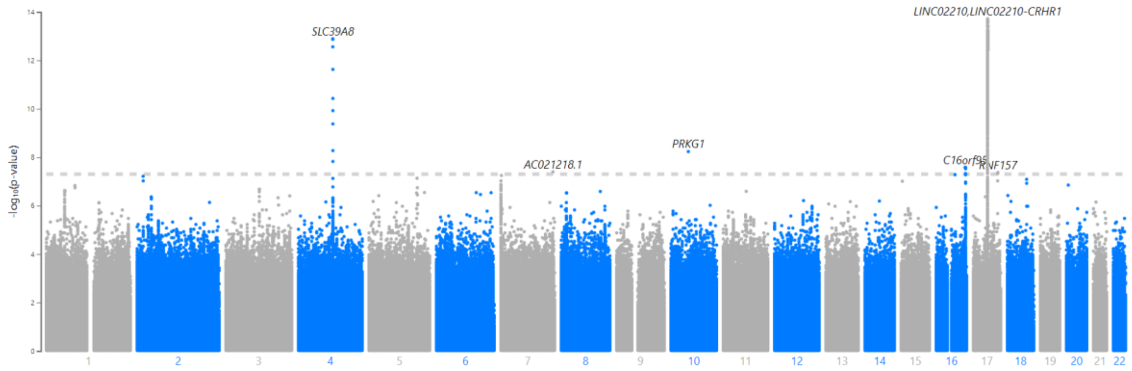


(c) SKAT-CommonRare results (gene-based test with WES data)

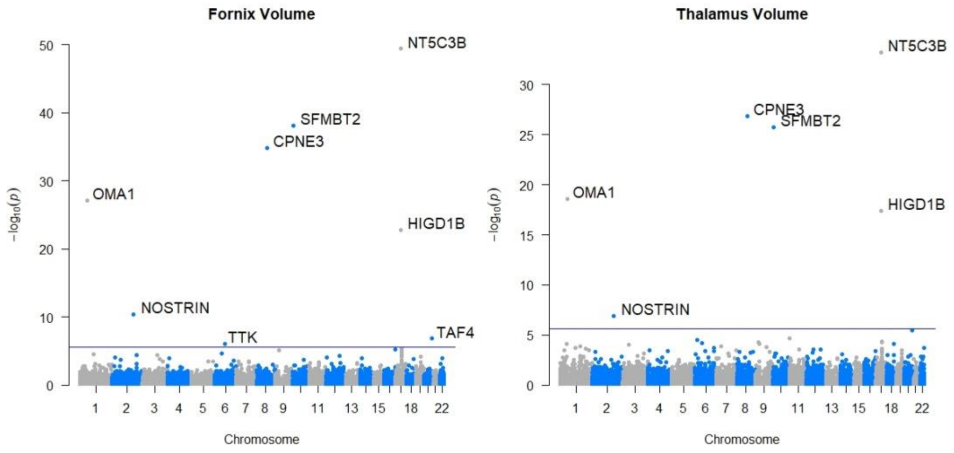


**Supplementary Figure 2** GWAS results on the average voxel values in the fornix and the lower part of the thalamus.

(a) Inverse variance weighting method

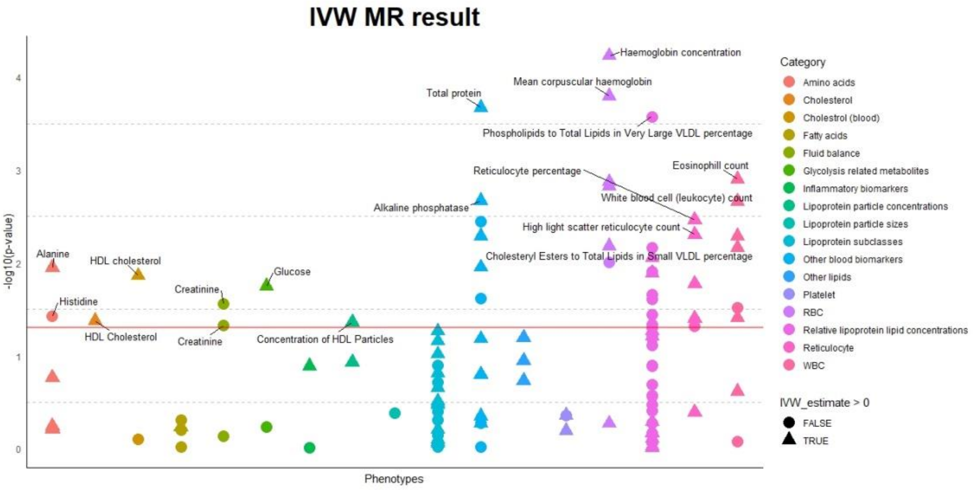


(b) Weighted median method

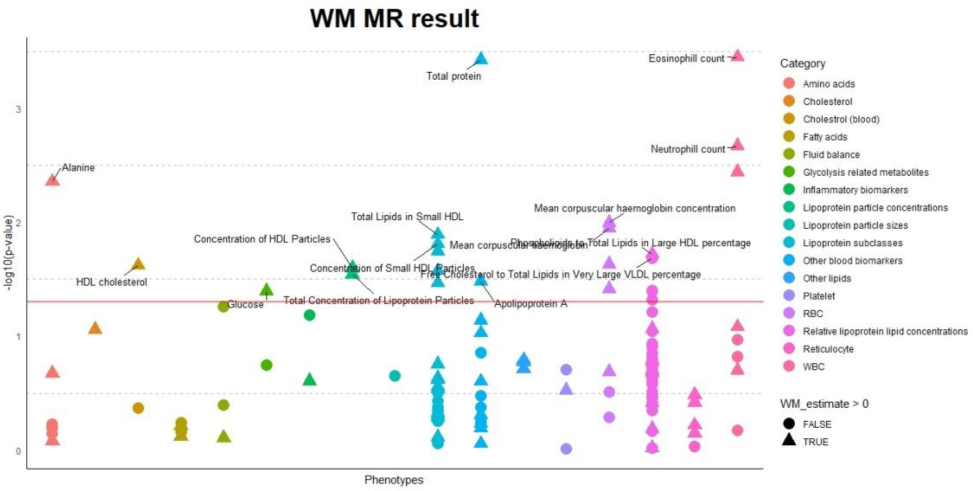


**Supplementary Figure 3** PheWAS plots of the linear MR causal estimates from (a) Inverse variance weighting method and (b) Weighted median method

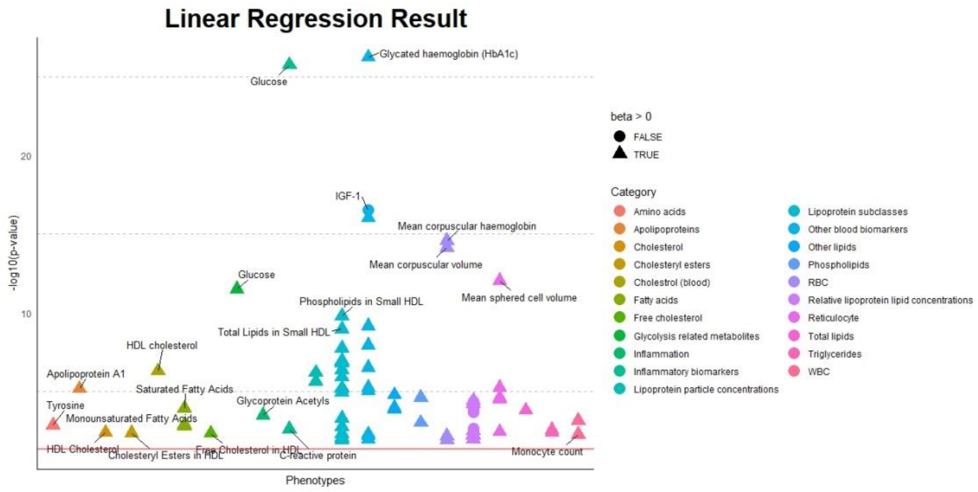


**Supplementary Figure 4** A plot of p-values of the linear regression coefficients (delta age ~ phenotype + sex + age + PC scores + cross-validation batch)

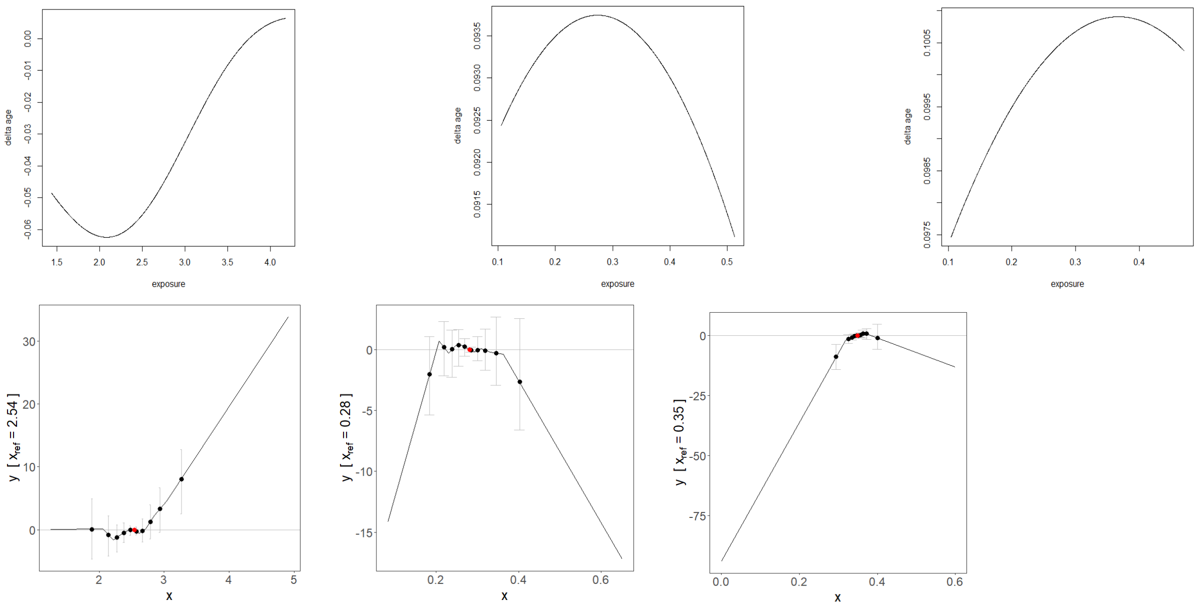


**Supplementary Figure 5** Plots of the nonlinear causal relationship between the biomarkers and delta age. The first row shows the plots with kernel IV regression, and the next row shows the plots with piecewise MR. The order of the plots is total cholines, total lipids in small LDL, and cholesteryl esters to total lipids in very large HDL percentage.

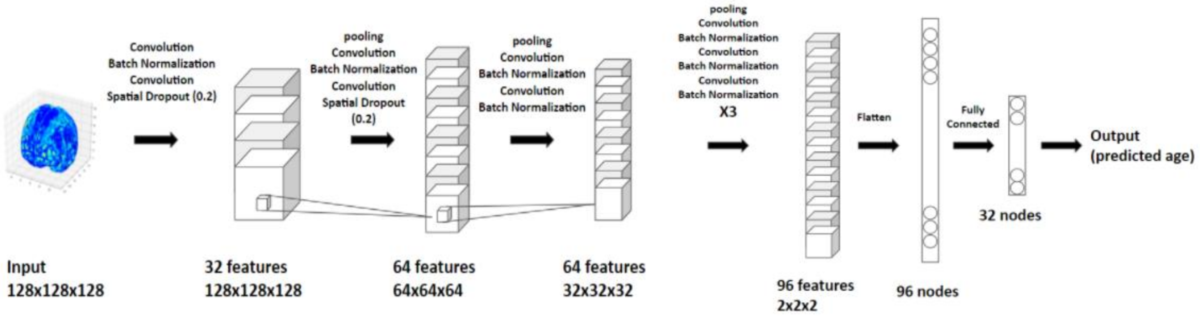


**Supplementary Figure 6** The convolutional neural network structure of the age prediction model

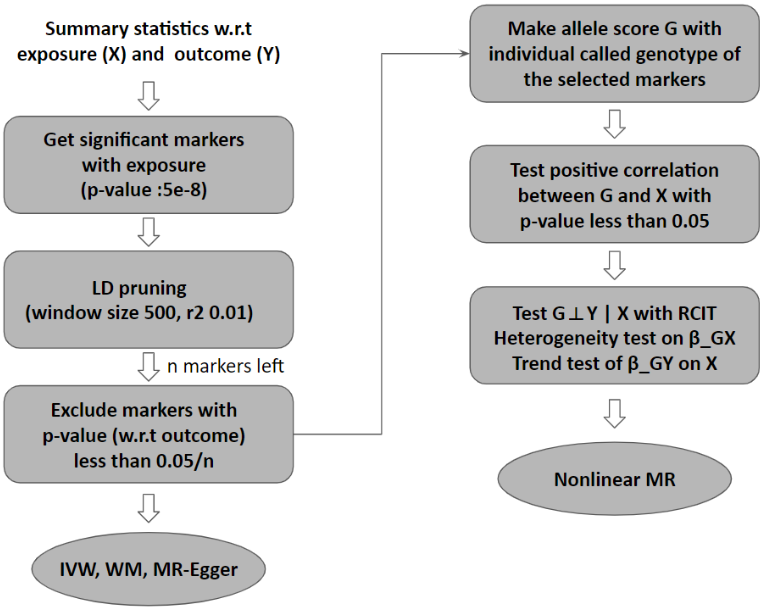


**Supplementary Figure 7** Workflow of the linear and nonlinear MR analysis

**Supplementary Table 1** SNPs in top loci associated with volume in (a) the fornix (b) the lower part of the thalamus (SAIGE, single-variant test with array-genotyped and imputed data)

(a)

| Chromosome | Location | rsID (GRCh37) | Nearest gene | Effect size | P-value |
|---|---|---|---|---|---|
| 4 | 103,198,082 | rs13135092 | SLC39A8 | -0.116 | 1.00E-18 |
| 17 | 44,364,584 | rs2696498 | ARL17B | -0.078 | 4.42E-16 |
| 2 | 198,905,270 | rs892514 | PLCL1 | -0.044 | 6.67E-09 |
| 16 | 87,227,001 | rs12921632 | C16orf95 | -0.045 | 1.18E-08 |
| 1 | 93,259,188 | rs561791307 | EVI5 | 1.619 | 1.31E-08 |
| 3 | 760,388 | rs543823118 | LINC01266 | 1.062 | 2.31E-08 |

(b)

| Chromosome | Location | rsID (GRCh37) | Nearest gene | Effect size | P-value |
|---|---|---|---|---|---|
| 17 | 43,707,848 | [illegible] | LINC02210, LINC02210-CRHR1 | -0.063 | 1.92E-14 |
| 4 | 103,198,082 | rs13135092 | SLC39A8 | -0.093 | 1.29E-13 |
| 10 | 52,871,149 | rs535001460 | PRKG1 | 1.552 | 5.85E-09 |
| 16 | 87,231,160 | rs4843555 | C16orf95 | -0.038 | 2.63E-08 |
| 7 | 155,766,573 | rs371967302 | AC021218.1 | -3.344 | 4.00E-08 |
| 17 | 74,151,456 | rs894544 | RNF157 | 0.060 | 4.25E-08 |

**Supplementary Table 2** Gene regions associated with volume in (a) the fornix (b) the lower part of the thalamus (SKAT-CommonRare, gene-based test with WES data)

(a)

| Chromosome | Starting location (GRCh38) | Gene | P-value |
|---|---|---|---|
| 17 | 41825424 | NT5C3B | 4.04E-50 |
| 10 | 7163670 | SFMBT2 | 8.24E-39 |
| 8 | 86528449 | CPNE3 | 1.39E-35 |
| 1 | 58480654 | OMA1 | 7.46E-28 |
| 17 | 44848057 | HIGD1B | 1.86E-23 |
| 2 | 168802560 | NOSTRIN | 4.62E-11 |
| 20 | 61976072 | TAF4 | 1.56E-07 |
| 6 | 80005748 | TTK | 9.43E-07 |

(b)

| Chromosome | Starting location (GRCh38) | Gene | P-value |
|---|---|---|---|
| 17 | 41825424 | NT5C3B | 6.25E-34 |
| 8 | 86528449 | CPNE3 | 1.37E-27 |
| 10 | 7163670 | SFMBT2 | 1.85E-26 |
| 1 | 58480654 | OMA1 | 2.53E-19 |
| 17 | 44848057 | HIGD1B | 3.72E-18 |
| 2 | 168802560 | NOSTRIN | 1.35E-07 |

**Supplementary Table 3** Genetic correlation and correlation coefficients among delta age and average voxel values of the fornix and the lower part of the thalamus

| Region | Genetic correlation | P-value | Correlation coefficient | P-value |
|---|---|---|---|---|
| Fornix | -0.320 | 4.00E-04 | -0.099 | 1.56E-74 |
| Thalamus | -0.503 | 3.19E-08 | -0.135 | 2.46E-138 |

**Supplementary Table 4** A list of causal estimates of 55 biomarkers that did not pass multiple testing correction in the MR-Egger regression

| Biomarker | Estimates | P-value |
|---|---|---|
| White blood cell (leukocyte) count | 0.100 | 8.01E-03 |
| Total Fatty Acids | 0.179 | 1.13E-02 |
| Monounsaturated Fatty Acids | 0.160 | 1.83E-02 |
| Lymphocyte percentage | -0.150 | 2.41E-02 |
| Glutamine | -0.209 | 3.05E-02 |
| Triglycerides in Small HDL | 0.146 | 3.06E-02 |
| Basophil percentage | -0.248 | 3.11E-02 |
| Mean corpuscular haemoglobin | 0.054 | 3.44E-02 |
| Cholesteryl Esters to Total Lipids in Small LDL percentage | 0.111 | 3.90E-02 |
| Phospholipids to Total Lipids in Very Large VLDL percentage | 0.191 | 3.98E-02 |
| Phospholipids to Total Lipids in Small LDL percentage | -0.121 | 3.98E-02 |
| C-reactive protein | -0.074 | 4.59E-02 |
| Testosterone | -0.189 | 4.86E-02 |
| Haemoglobin concentration | 0.096 | 7.99E-02 |
| Mean corpuscular volume | 0.047 | 1.17E-01 |
| Concentration of Small HDL Particles | 0.148 | 2.03E-01 |
| Alkaline phosphatase | -0.039 | 2.06E-01 |
| Phospholipids to Total Lipids in Large HDL percentage | -0.090 | 2.46E-01 |
| Free Cholesterol to Total Lipids in Very Large VLDL percentage | -0.112 | 2.64E-01 |
| Creatinine | -0.090 | 2.70E-01 |
| Mean corpuscular haemoglobin concentration | 0.101 | 3.46E-01 |
| High light scatter reticulocyte percentage | -0.047 | 3.92E-01 |
| Cholesteryl Esters in Small HDL | 0.096 | 3.95E-01 |
| Histidine | -0.138 | 4.07E-01 |
| HDL cholesterol | 0.038 | 4.12E-01 |
| Cholesteryl Esters to Total Lipids in Small VLDL percentage | -0.048 | 4.36E-01 |
| Total Lipids in Small HDL | 0.083 | 4.66E-01 |
| Phospholipids in Medium HDL | -0.063 | 4.77E-01 |
| Alanine | 0.102 | 4.81E-01 |
| Gamma glutamyltransferase | 0.038 | 4.88E-01 |
| Free Cholesterol to Total Lipids in Small VLDL percentage | 0.028 | 5.19E-01 |
| Total Concentration of Lipoprotein Particles | 0.071 | 5.30E-01 |
| Glucose | 0.158 | 5.40E-01 |
| Reticulocyte percentage | -0.030 | 5.70E-01 |
| IGF-1 | 0.029 | 5.72E-01 |
| Monocyte count | 0.022 | 5.72E-01 |
| Phospholipids to Total Lipids in Small VLDL percentage | 0.024 | 5.82E-01 |
| High light scatter reticulocyte count | -0.025 | 6.68E-01 |
| Apolipoprotein A | 0.021 | 6.69E-01 |
| Creatinine | -0.087 | 6.95E-01 |
| Red blood cell (erythrocyte) distribution width | 0.012 | 7.59E-01 |
| Cholesteryl Esters to Total Lipids in Large VLDL | -0.023 | 7.75E-01 |

| | | |
|---|---|---|
| percentage | | |
| Cholesterol to Total Lipids in Large VLDL percentage | 0.024 | 7.91E-01 |
| Cholesterol to Total Lipids in Small VLDL percentage | -0.015 | 7.95E-01 |
| HDL Cholesterol | -0.013 | 8.63E-01 |
| Triglycerides to Total Lipids in Small VLDL percentage | 0.009 | 8.67E-01 |
| Phospholipids in Small HDL | 0.016 | 8.74E-01 |
| Haematocrit percentage | 0.012 | 8.74E-01 |
| Free Cholesterol to Total Lipids in Very Small VLDL percentage | 0.013 | 8.80E-01 |
| Aspartate aminotransferase | 0.008 | 8.83E-01 |
| Reticulocyte count | 0.008 | 8.86E-01 |
| Cholesterol to Total Lipids in Medium VLDL percentage | -0.007 | 8.92E-01 |
| Concentration of HDL Particles | 0.008 | 9.29E-01 |
| Mean reticulocyte volume | 0.002 | 9.62E-01 |
| Cholesteryl Esters to Total Lipids in Medium VLDL percentage | 0.001 | 9.86E-01 |

**Supplementary Table 5** A list of causal estimates and p-values of 59 biomarkers in the order of weighted median p-value (Bolded p-values mean that they passed multiple testing corrections)

| **Biomarker** | **Est (WM)** | **P (WM)** | **Est (IVW)** | **P (IVW)** |
|---|---|---|---|---|
| Eosinophil count | 0.080 | **3.58E-04** | 0.049 | **1.26E-03** |
| Total protein | 0.104 | **3.77E-04** | 0.076 | **2.12E-04** |
| Neutrophil count | 0.094 | **2.16E-03** | 0.058 | **6.83E-03** |
| Eosinophil percentage | 0.073 | 3.63E-03 | 0.035 | 3.90E-02 |
| Alanine | 0.183 | 4.41E-03 | 0.114 | 1.13E-02 |
| Mean corpuscular haemoglobin concentration | 0.143 | 1.01E-02 | 0.129 | **1.49E-03** |
| Mean corpuscular haemoglobin | 0.059 | 1.13E-02 | 0.051 | **1.60E-04** |
| Total Lipids in Small HDL | 0.142 | 1.29E-02 | 0.071 | 9.61E-02 |
| Concentration of Small HDL Particles | 0.142 | 1.56E-02 | 0.044 | 3.08E-01 |
| Phospholipids in Small HDL | 0.114 | 1.80E-02 | 0.037 | 3.22E-01 |
| Phospholipids to Total Lipids in Large HDL percentage | 0.098 | 1.90E-02 | 0.075 | 1.29E-02 |
| Free Cholesterol to Total Lipids in Very Large VLDL percentage | -0.105 | 2.08E-02 | -0.052 | 1.29E-01 |
| Haemoglobin concentration | 0.083 | 2.33E-02 | 0.091 | **5.89E-05** |
| HDL cholesterol | 0.069 | 2.40E-02 | 0.047 | **1.37E-02** |
| Concentration of HDL Particles | 0.108 | 2.53E-02 | 0.067 | 4.36E-02 |
| Phospholipids in Medium HDL | 0.098 | 2.68E-02 | 0.041 | 2.19E-01 |
| Total Concentration of Lipoprotein Particles | 0.112 | 2.86E-02 | 0.063 | 1.18E-01 |
| Apolipoprotein A | 0.069 | 3.29E-02 | 0.038 | 6.55E-02 |
| Cholesteryl Esters in Small HDL | 0.119 | 3.42E-02 | 0.080 | 5.41E-02 |
| Mean corpuscular volume | 0.046 | 3.85E-02 | 0.046 | **1.34E-03** |
| Cholesteryl Esters to Total Lipids in Large VLDL percentage | -0.088 | 4.01E-02 | -0.070 | 2.47E-02 |
| Glucose | 0.175 | 4.02E-02 | 0.140 | 1.77E-02 |
| Cholesterol to Total Lipids in Large VLDL percentage | -0.093 | 4.04E-02 | -0.065 | 6.26E-02 |
| Cholesteryl Esters to Total Lipids in Small VLDL percentage | -0.079 | 4.76E-02 | -0.071 | 6.82E-03 |
| Creatinine | -0.065 | 5.54E-02 | -0.054 | 2.76E-02 |
| Phospholipids to Total Lipids in Very Large VLDL percentage | -0.082 | 6.10E-02 | -0.126 | 2.66E-04 |
| C-reactive protein | -0.064 | 6.50E-02 | -0.001 | 9.78E-01 |
| White blood cell (leukocyte) count | 0.047 | 8.30E-02 | 0.056 | **2.18E-03** |
| HDL Cholesterol | 0.077 | 8.75E-02 | 0.059 | 4.16E-02 |
| Lymphocyte percentage | -0.048 | 1.07E-01 | -0.048 | 3.00E-02 |
| Free Cholesterol to Total Lipids in Very Small VLDL percentage | -0.073 | 1.16E-01 | -0.068 | 4.70E-02 |
| Cholesterol to Total Lipids in Small VLDL percentage | -0.063 | 1.23E-01 | -0.070 | 1.24E-02 |
| Testosterone | -0.129 | 1.39E-01 | -0.124 | 2.41E-02 |

| | | | | |
|---|---|---|---|---|
| Basophil percentage | -0.083 | 1.50E-01 | -0.008 | 8.42E-01 |
| Cholesterol to Total Lipids in Medium VLDL percentage | -0.056 | 1.55E-01 | -0.055 | 2.21E-02 |
| Cholesteryl Esters to Total Lipids in Medium VLDL percentage | -0.055 | 1.64E-01 | -0.048 | 4.58E-02 |
| Triglycerides to Total Lipids in Small VLDL percentage | 0.055 | 1.70E-01 | 0.070 | 8.71E-03 |
| Monocyte count | 0.032 | 2.00E-01 | 0.043 | **5.12E-03** |
| Haematocrit percentage | 0.047 | 2.06E-01 | 0.072 | **6.59E-03** |
| Gamma glutamyltransferase | 0.035 | 2.50E-01 | 0.060 | **5.15E-03** |
| Phospholipids to Total Lipids in Small VLDL percentage | -0.043 | 2.50E-01 | -0.050 | 3.58E-02 |
| Free Cholesterol to Total Lipids in Small VLDL percentage | -0.043 | 2.65E-01 | -0.047 | 4.96E-02 |
| High light scatter reticulocyte count | 0.027 | 3.29E-01 | 0.056 | **4.94E-03** |
| IGF-1 | -0.025 | 3.34E-01 | -0.054 | **3.56E-03** |
| Cholesteryl Esters to Total Lipids in Small LDL percentage | 0.037 | 3.82E-01 | 0.008 | 7.73E-01 |
| High light scatter reticulocyte percentage | 0.023 | 3.87E-01 | 0.047 | 1.68E-02 |
| Creatinine | -0.054 | 4.03E-01 | -0.104 | 4.69E-02 |
| Phospholipids to Total Lipids in Small LDL percentage | -0.037 | 4.06E-01 | 0.002 | 9.45E-01 |
| Triglycerides in Small HDL | 0.029 | 4.95E-01 | -0.012 | 6.81E-01 |
| Red blood cell (erythrocyte) distribution width | -0.018 | 5.15E-01 | -0.043 | **9.75E-03** |
| Aspartate aminotransferase | -0.017 | 5.65E-01 | 0.053 | **1.11E-02** |
| Reticulocyte percentage | 0.014 | 6.07E-01 | 0.058 | **3.43E-03** |
| Total Fatty Acids | -0.017 | 6.89E-01 | -0.019 | 4.86E-01 |
| Histidine | -0.031 | 7.12E-01 | -0.124 | 3.74E-02 |
| Reticulocyte count | 0.010 | 7.15E-01 | 0.040 | 3.94E-02 |
| Monounsaturated Fatty Acids | 0.013 | 7.58E-01 | 0.015 | 5.87E-01 |
| Glutamine | 0.012 | 8.34E-01 | 0.054 | 1.72E-01 |
| Alkaline phosphatase | 0.004 | 8.79E-01 | 0.049 | **2.14E-03** |
| Mean reticulocyte volume | -0.002 | 9.27E-01 | -0.035 | 4.78E-02 |

**Supplementary Table 6** The Linear regression coefficients and p-values of significant phenotypes after Benjamini-Hochberg procedure (delta age ~ phenotype + covariates) (a) blood-related phenotypes (b) metabolomic biomarkers

(a)

| Biomarker | Estimates | P-value |
|---|---|---|
| Glycated haemoglobin (HbA1c) | 0.034 | 7.31E-28 |
| Glucose | 0.172 | 1.11E-26 |
| IGF-1 | -0.025 | 5.61E-18 |
| Gamma glutamyltransferase | 0.004 | 2.45E-17 |
| Mean corpuscular haemoglobin | 0.073 | 1.43E-15 |
| Mean corpuscular volume | 0.029 | 4.53E-15 |
| Mean sphered cell volume | 0.023 | 6.76E-13 |
| Apolipoprotein A | 0.399 | 8.02E-11 |
| Aspartate aminotransferase | 0.010 | 6.18E-09 |
| HDL cholesterol | 0.242 | 7.51E-08 |
| Alanine aminotransferase | 0.006 | 1.27E-07 |
| Alkaline phosphatase | 0.003 | 4.82E-06 |
| High light scatter reticulocyte percentage | 0.339 | 5.96E-06 |
| Calcium | 0.754 | 9.03E-06 |
| Immature reticulocyte fraction | 1.103 | 2.41E-05 |
| High light scatter reticulocyte count | 7.092 | 2.80E-05 |
| Haemoglobin concentration | 0.056 | 5.81E-04 |
| White blood cell (leukocyte) count | 0.029 | 7.55E-04 |
| Urate | 0.001 | 1.46E-03 |
| C-reactive protein | 0.013 | 2.60E-03 |
| Mean reticulocyte volume | 0.006 | 3.65E-03 |
| Phosphate | 0.288 | 4.25E-03 |
| Monocyte count | 0.240 | 4.76E-03 |
| Lymphocyte count | 0.045 | 5.86E-03 |

| | | |
|---|---|---|
| Albumin | 0.016 | 1.05E-02 |
| Red blood cell (erythrocyte) count | -0.116 | 1.11E-02 |
| Mean corpuscular haemoglobin concentration | 0.038 | 1.16E-02 |
| Cystatin C | 0.318 | 1.51E-02 |
| Haematocrit percentage | 0.013 | 1.91E-02 |
| Creatinine | -0.003 | 1.91E-02 |

(b)

| Biomarker | Estimates | P-value |
|---|---|---|
| Glucose | 0.227 | 2.36E-12 |
| Phospholipids in Small HDL | 2.316 | 1.27E-10 |
| Total Lipids in Small HDL | 1.279 | 1.04E-09 |
| Phospholipids in Medium HDL | 2.153 | 1.36E-09 |
| Total Lipids in Medium HDL | 0.908 | 1.04E-08 |
| Free Cholesterol in Small HDL | 11.162 | 7.91E-08 |
| Cholesterol in Small HDL | 2.849 | 1.08E-07 |
| Concentration of HDL Particles | 76.645 | 1.09E-07 |
| Concentration of Medium HDL Particles | 203.986 | 1.19E-07 |
| Concentration of Small HDL Particles | 130.551 | 1.69E-07 |
| Cholesteryl Esters in Small HDL | 3.474 | 4.56E-07 |
| Total Concentration of Lipoprotein Particles | 69.391 | 5.10E-07 |
| Apolipoprotein A1 | 0.729 | 7.47E-07 |
| Cholesterol in Medium HDL | 1.435 | 7.71E-07 |
| Cholesteryl Esters in Medium HDL | 1.780 | 7.98E-07 |
| Free Cholesterol in Medium HDL | 7.036 | 1.27E-06 |
| Phospholipids in HDL | 0.532 | 2.44E-06 |
| Phosphoglycerides | 0.400 | 5.33E-06 |
| Phospholipids to Total Lipids in Large HDL percentage | 5.010 | 2.23E-05 |

| | | |
|---|---|---|
| Total Lipids in HDL | 0.237 | 2.34E-05 |
| Linoleic Acid to Total Fatty Acids percentage | -4.065 | 2.91E-05 |
| Phosphatidylcholines | 0.389 | 3.58E-05 |
| Total Cholines | 0.347 | 5.15E-05 |
| Saturated Fatty Acids to Total Fatty Acids percentage | 6.687 | 5.94E-05 |
| Polyunsaturated Fatty Acids to Total Fatty Acids percentage | -3.498 | 8.70E-05 |
| Saturated Fatty Acids | 0.136 | 1.11E-04 |
| Omega-6 Fatty Acids to Total Fatty Acids percentage | -3.506 | 1.53E-04 |
| Glycoprotein Acetyls | 1.016 | 3.18E-04 |
| Triglycerides in Medium HDL | 6.092 | 5.00E-04 |
| Total Phospholipids in Lipoprotein Particles | 0.255 | 5.47E-04 |
| Monounsaturated Fatty Acids | 0.130 | 1.24E-03 |
| HDL Cholesterol | 0.353 | 1.29E-03 |
| Free Cholesterol in HDL | 1.570 | 1.40E-03 |
| Tyrosine | 7.450 | 1.41E-03 |
| Cholesteryl Esters in HDL | 0.446 | 1.42E-03 |
| Total Fatty Acids | 0.043 | 1.58E-03 |
| Apolipoprotein B to Apolipoprotein A1 ratio | -0.626 | 1.62E-03 |
| Triglycerides in Large LDL | 4.120 | 1.63E-03 |
| Triglycerides in HDL | 2.107 | 2.72E-03 |
| Polyunsaturated Fatty Acids to Monounsaturated Fatty Acids ratio | -0.280 | 2.88E-03 |
| Free Cholesterol to Total Lipids in Very Small VLDL percentage | -16.753 | 2.97E-03 |
| Triglycerides to Total Lipids in Large LDL percentage | 6.003 | 3.01E-03 |
| Triglycerides in LDL | 2.364 | 3.76E-03 |
| Phospholipids to Total Lipids in Small HDL percentage | 7.892 | 3.82E-03 |
| Triglycerides in IDL | 3.623 | 4.80E-03 |

| | | |
|---|---|---|
| Cholesterol to Total Lipids in Large LDL percentage | -5.665 | 5.27E-03 |
| Triglycerides to Total Lipids in Medium LDL percentage | 6.145 | 6.34E-03 |
| Phospholipids to Total Lipids in IDL percentage | -9.559 | 7.64E-03 |
| Triglycerides in Medium LDL | 8.653 | 7.69E-03 |
| Triglycerides to Total Lipids in Very Small VLDL percentage | 2.280 | 8.20E-03 |
| Triglycerides in Small HDL | 4.897 | 1.01E-02 |
